# Two-Stage Machine Learning Based Prediction of Thrombophilia Management

**DOI:** 10.1101/2025.10.22.25338547

**Authors:** Hannah L. McRae, Fabian Kahl, Maximilian Kapsecker, Heiko Rühl, Stephan M. Jonas, Bernd Pötzsch

## Abstract

Thrombophilia diagnosis and management rely on the nuanced interpretation of clinical history, risk factors, and laboratory data, yet significant variability exists in clinical practice due to subjective assessment, institutional differences, and limited consensus guidelines. In this retrospective study, we evaluated the use of a two-stage machine learning (ML) approach to predict thrombophilia diagnosis and subsequent anticoagulation treatment. The study included data (14 clinical and 26 laboratory features) from 496 patients evaluated at the University Hospital Bonn between 2019 and 2024. The hybrid nature of the diagnostic categories combining ordinal (including thrombophilia diagnosis severity and treatment recommendations) and categorical (e.g. antiphospholipid syndrome) classes necessitated a two-stage approach. XGBClassifier was used to distinguish categorical from ordinal classes, followed by an ordinal-specific XGBOrdinalV2 model. A sliding window approach improved classification performance across all ordinal categories reaching sensitivities above 89% across all ordinal classes. Feature importance analysis revealed that age at first thrombosis and antiphospholipid antibody status were key predictive variables. Of the 496 patients, 362 (73%) experienced no discrepancy between the ML-based predictions and the practitioner diagnosis and subsequent treatment recommendations. Re-evaluation of the remaining 134 patients revealed that, while the ML models correctly classified 36 patients, it underestimated or overestimated the thrombophilia severity in 71 and 27 patients, respectively. This study highlights the potential of interpretable ML models to support standardized thrombophilia management and improve diagnostic accuracy. Future prospective studies and external validation are needed to assess generalizability and clinical impact.

## Introduction

Thrombophilia is characterized by a predisposition to thrombosis formation and can be caused by both hereditary and acquired conditions [1]. The diagnosis of thrombophilia is made on the basis of a comprehensive collection and analysis of clinical and laboratory data. Relevant clinical data include the number and localization of thrombotic events, age at the first episode of thrombosis, risk factor(s) present at the time of thrombosis to determine whether the thrombosis was provoked or unprovoked, and family history of thrombosis or thrombophilia. In addition, some thrombophilias are associated with miscarriage and other pregnancy complications [2]. Utilizing these data, the individual probability of thrombosis can be calculated, as validated by several probability models developed over the past two decades [3-6].

Laboratory screening for hereditary and acquired thrombophilia involves a combination of standard and specialized coagulation assays, functional tests, and molecular genetic testing for factor V Leiden (FVL) and prothrombin G20210A (F2G20210A) mutation, antithrombin (AT), protein C (PC), and protein S (PS) deficiency, as well as antiphospholipid antibody (aPL) testing including lupus anticoagulant (LA), anticardiolipin antibody (aCL), and beta-2 glycoprotein 1 antibody (ß2GP1). While not part of routine thrombophilia workups, testing for other acquired thrombophilic conditions such as heparin-induced thrombocytopenia (HIT), paroxysmal nocturnal hemoglobinuria (PNH), and the somatic JAK2 mutation, among others [7-9], may also be performed depending on the clinical scenario.

Thrombophilia management is highly individualized and can be challenging, especially for providers who are not specialized in hemostasis and thrombosis. Several factors such as differences in institutional protocols and guidelines, patient insurance coverage, and individual practice philosophies, which are inherently subjective, can result in variability in diagnosis and management. This leaves an opportunity for optimization, and the implementation of artificial intelligence (AI)- and machine learning (ML)-powered solutions could potentially increase standardization and improve patient care in thrombophilia management.

In recent years, ML has emerged as a promising tool for medical decision-making as it enables the efficient analysis of large volumes of high-dimensional data. In particular, ML makes it possible to emulate human clinical decision-making, extrapolate new information, and optimize clinical protocols, thus potentially improving clinical practice and diagnostic accuracy over time [10]. Moreover, recent studies demonstrate that tree-based models such as XGBoost or Random Forests are still superior to novel deep learning methods for the analysis of tabular datasets [11]. In a 2023 systematic review and pooled analysis, Chiasakul et al. evaluated the effectiveness of various AI models in predicting and diagnosing venous thromboembolism (VTE) in different clinical settings. The authors reviewed 745 titles and abstracts, ultimately including 20 studies [12]. Of these, only two papers mentioned thrombophilia [13, 14], and both did not investigate patients with thrombophilia exclusively, but rather included thrombophilia as an additional feature in their ML models. To the best of our knowledge, no previous research to date has explored the use of ML tools for thrombophilia management.

This study aimed to develop predictive machine learning models for thrombophilia diagnosis and risk-adjusted anticoagulant treatment among patients undergoing thrombophilia evaluation. While treatment recommendations were entirely ordinal, ranging from no treatment to the recommendation of indefinite oral anticoagulation, the diagnosis categories were a mix of categorical and ordinal classes. A two-stage approach to the ML model design was implemented in order to account for the presence of both categorical and ordinal classes in the dataset. In the first stage, the program XGBClassifier, an implementation of XGBoost for classification tasks [15], predicted whether a patient belonged to a categorical or ordinal class for diagnosis and VTE risk assessment. If a patient was in any ordinal class, the second stage used XGBOrdinalV2, an implementation of XGBoost for ordinal classification tasks [16], to predict the specific ordinal class. For the prediction of treatment, only XGBOrdinalV2 was used. The use of XGBoost is well-established in medicine and medical research and is used for predicting various diseases and disease states [17-19].

The results of our study demonstrate that the implementation of ML tools shows promise for effectively supporting the complex decision-making process in clinical management of patients with thrombophilia.

## Methods

### Clinical and Laboratory Data Collection

This single-center, retrospective study was performed at the Institute of Experimental Hematology and Transfusion Medicine at the University Hospital Bonn, Germany. Data sets of patients who completed thrombophilia evaluation between November 2019 and August 2024 were available for the study. Patients who had VTE within six weeks prior to blood collection, arterial thrombosis, as well as conditions associated with thromboembolism such as malignancy, beta thalassemia, cardiovascular disease, and May-Thurner syndrome were excluded from analysis.

The data collected from the electronic medical record (EMR) included age and sex, body mass index (BMI), history of thrombosis including the number of thrombotic events, thrombosis localization, provoking factors at time of thrombosis, age at first thrombosis (if applicable), family history of thrombosis (if applicable), any history of miscarriage (in females), concomitant medical conditions, and any current anticoagulation or antiplatelet therapy including dosage. These data are henceforth referred to as clinical data.

The results of all relevant laboratory tests were also recorded including basic hemostasis parameters: platelet count, prothrombin time (PT), activated partial thromboplastin time (aPTT), fibrinogen; thrombophilic risk factors: factor VIII, factor Xa- or thrombin-based antithrombin activity (AT-FXa/FIIa), protein C (PC), free protein S (PS), activated protein C resistance (APCR), aPL including lupus anticoagulant (LA), anticardiolipin antibody (aCL IgG and IgM), beta-2 glycoprotein 1 antibody (anti-ß2GP1 IgG and IgM), anti-phosphatidylserine prothrombin antibodies (aPSPT IgG and IgM), factor V Leiden (FVL) mutation including HR_2_ haplotype, and prothrombin G20210A (F2G20210A) mutation; and procoagulant biomarkers: prothrombin fragment 1+2 (F 1+2) and D-dimer. These data are henceforth referred to as laboratory data. Laboratory testing was performed locally as previously described [20]. Of note, for patients taking direct oral anticoagulants (DOAC), blood collection was performed 12 hours (apixaban, dabigatran) or 24 hours (edoxaban, rivaroxaban) following the most recent dose. Patients taking vitamin K antagonists (VKA) were tested when they were switched to low molecular weight heparin (LMWH) for medical reasons such as tooth extraction or other planned procedures. In addition, DOAC and LMWH levels were measured at the time of testing to ensure that there was no clinically significant interference on thrombophilia test results.

Data including final thrombophilia diagnosis and treatment decisions were additionally recorded for comparison at the end of the study. The Ethics Committee of the University of Bonn Faculty of Medicine sanctioned the collection of data from the EMR and waived the requirement for written informed consent due to the retrospective nature of the study. The study was performed in accordance with the Declaration of Helsinki. All authors had access to primary data.

### Stratification of Patient Cohort by Diagnosis, VTE Risk, and Treatment

Thrombophilia diagnoses were made and subsequent anticoagulant treatment after completion of full-intensity primary anticoagulation were recommended as previously described (Table 1) [20]. No anticoagulant treatment, except standard of care thromboprophylaxis in high-risk situations, is recommended for patients in whom thrombophilia has been ruled out. For patients with low-risk diagnoses, routine prophylaxis extended to low-risk situations such as long-distance travel or bed rest for more than 3 days was recommended. Patients diagnosed with intermediate thrombophilia were recommended low-dose DOAC therapy for a prolonged or indefinite period, whereas those with high-risk thrombophilia were recommended standard-dose DOAC therapy for an indefinite period. Indefinite anticoagulation was defined as therapy without a defined endpoint, with reassessment of the risks and benefits every two to five years [20-25]. Patients with confirmed APS according to the revised Sapporo diagnostic criteria [26] were considered for indefinite VKA therapy with a therapeutic INR of 2-3.0 [27].

**Table 1.** Treatment recommendations based on thrombophilia diagnosis.

| Diagnosis | Treatment Recommendation |
| --- | --- |
| **D0:** Thrombophilia excluded | **T0:** standard of care prophylaxis in high-risk situations |
| **D1:** Low-risk thrombophilia | **T1:** Routine prophylaxis extended to low-risk situations |
| **D2:** Intermediate-risk thrombophilia | **T2:** Low-dose DOAC extended<br>**T3:** Low-dose DOAC indefinite |
| **D3:** High-risk thrombophilia | **T4:** Standard-dose DOAC indefinite |
| **D4:** Antiphospholipid syndrome | **T5:** VKA anticoagulation |

Of note, the final treatment recommendation was given at the discretion of the treating physician after consideration of all factors including personal and family history, physical examination, and all other relevant clinical and laboratory data.

### Model Architecture

The categories within the two target features for this study represented both ordinal and categorical data, necessitating the implementation of a two-stage approach to the model design (Supplemental Figure S1). Due to the hybrid categorical and ordinal nature of the diagnosis target feature, we used a two-stage approach (Supplemental Figure S1). For categorical classification, an XGBClassifier model [15] was used to predict whether the patient belongs to the categorical class or any ordinal class. For ordinal classification, an XGBOrdinalV2 model [16] was used to determine the exact ordinal class for patients identified as ordinal.

The dataset was split into train and test sets (80% and 20% respectively). We used the test sets to evaluate model performances. To prevent the occurrence of a randomly well-performing or badly-performing model, we conducted the experiment 100 times with 100 different random train and test splits. Additional technical details are outlined in the supplemental material.

### Statistical Analysis

To evaluate model performances, mean sensitivity, specificity, and precision ± standard deviation (std) per target feature were calculated for the 100 runs (the two-stage models for diagnosis and the ordinal models for treatment). These three metrics were analyzed with and without a one-class sliding window in the ordinal classes. In the sliding window approach, predictions were considered correct if they fell within one ordinal class of the actual value. Mean ± std confusion matrices were generated per target feature to assess classification performance across the two-stage models (used for diagnosis) and the ordinal models (used for treatment). To evaluate feature importance, the absolute mean ± std SHAP values [28] were calculated for each feature per target feature, stage, and class with values expressed in probability. Features with higher SHAP values had a greater overall influence on the model’s predicted class probabilities.

## Results

### Study population characteristics

Clinical and laboratory data from 496 patients (309 females) with a mean age of 46.4 years (median 47 years, range 1.5 months – 89 years) were included in the study. Table 2 summarizes the diagnosis of thrombophilia, clinical and laboratory data, and treatment decisions. 347 patients had a history of thrombosis including 214 DVT, 140 PE, 13 cerebral venous sinus thrombosis, 11 retinal vein thrombosis, 1 splanchnic vein thrombosis, and 13 portal vein thrombosis. 388 (78.2 %) patients were on antithrombotic treatment including 104 (21.0 %) rivaroxaban, 86 (17.3 %) apixaban, 16 (3.2 %) edoxaban, 6 (1.2 %) fondaparinux, 30 (6.0 %) enoxaparin, 6 (1.2 %) dabigatran, 31 (6.3 %) VKA, 50 (10.1 %) aspirin, 9 (1.8 %) clopidogrel.

**Table 2:** Stratification of total patient cohort by diagnosis. Thrombophilia diagnosis categories as follows: thrombophilia ruled out (D0); low-risk thrombophilia diagnosis (D1); intermediate-risk thrombophilia diagnosis (D2); high-risk thrombophilia diagnosis (D3); confirmed antiphospholipid syndrome (D4).

| Patient Characteristics | Total<br>(n=496) | D0<br>(n=112,<br>22.6%) | D1<br>(n=104,<br>20.8%) | D2<br>(n=226,<br>45.6%) | D3<br>(n=22,<br>4.4%) | D4<br>(n=32,<br>6.5%) |
| --- | --- | --- | --- | --- | --- | --- |
| Female, n (%) | 309<br>(62.3) | 82 (72.6) | 76 (73.1) | 125 (55.3) | 9 (36.0) | 19 (57.6) |
| Age, mean (range) | 46.4 (1.5<br>months-89<br>years) | 38.8 (1.5<br>months-85 years) | 41.8 (10-89 years) | 51.4 (2.4<br>months-84 years) | 51.4 (14-72 years) | 48.4 (6-72 years) |
| Total VTE events, n | 478 | 19 | 62 | 310 | 49 | 38 |
| VTE per patient, mean<br>(range) | 0.96 (0-6) | 0.17 (0-1) | 0.60 (0-3) | 1.37 (1-4) | 2.23 (1-6) | 1.19 (0-4) |
| Thereof spontaneous, n<br>(mean per patient) | 163 (0.3) | 2 (0.0) | 9 (0.1) | 115 (0.5) | 15 (0.7) | 22 (0.7) |
| Thereof in the setting of<br>mild risk factor(s), n<br>(mean per patient) | 143 (0.3) | 4 (0.0) | 30 (0.3) | 95 (0.4) | 6 (0.3) | 8 (0.3) |
| Thereof provoked by<br>major risk factor(s), n<br>(mean per patient) | 31 (0.1) | 8 (0.1) | 10 (0.1) | 13 (0.1) | 0 (0.0) | 0 (0.0) |
| Age at first VTE, mean<br>(range) | 46.1 (11<br>days-87<br>years) | 48.3 (2.4<br>months-79 years) | 45.5 (20-87 years) | 46.7 (1-82<br>years) | 39.4 (1-68 years) | 46.1 (3-72 years) |
| Het./hom. FVL, n (%) | 89 (17.9) | 2 (1.8) | 32 (30.8) | 39 (17.3) | 11 (50.0) | 5 (15.6) |
| Het./hom. F2-G20210A, n (%) | 23 (4.6) | 0 (0.0) | 10 (9.6) | 11 (4.9) | 2 (9.1) | 0 (0.0) |
| Combined het. FVL/F2-G20210A, n (%) | 4 (0.8) | 0 (0.0) | 2 (1.9) | 1 (0.4) | 1 (4.5) | 0 (0.0) |
| AT deficiency, n (%) | 5 (1.0) | 0 (0.0) | 2 (1.9) | 1 (0.4) | 2 (9.1) | 0 (0.0) |
| PC deficiency, n (%) | 7 (1.4) | 0 (0.0) | 5 (4.8) | 0 (0.0) | 2 (9.1) | 0 (0.0) |
| PS deficiency, n (%) | 10 (2.0) | 0 (0.0) | 5 (4.8) | 1 (0.4) | 4 (18.2) | 0 (0.0) |
| Triple positive APS, n (%) | 30 (6.0) | 0 (0.0) | 5 (4.8) | 2 (0.9) | 2 (9.1) | 21 (65.6) |
| LA positive, n (%) | 46 (9.3) | 3 (2.7) | 8 (7.7) | 10 (4.4) | 1 (4.5) | 24 (75.0) |
| aPL positive but LA negative, n (%) | 58 (11.7) | 16 (14.3) | 12 (11.5) | 19 (8.4) | 3 (13.6) | 8 (25.0) |
| Heterozygous FVL and triple positive APS, n (%) | 5 (1.0) | 0 (0.0) | 1 (1.0) | 1 (0.4) | 1 (4.5) | 2 (6.3) |
| None, n (%) | 274 (55.2) | 92 (82.1) | 37 (35.6) | 140 (61.9) | 5 (22.7) | 0 (0.0) |
| T0, n (%) | 112 (22.6) | 112 (100) | 0 (0) | 0 (0) | 0 (0) | 0 (0) |
| T1, n (%) | 108 (21.8) | 0 (0) | 104 (100) | 1 (0.4) | 0 (0) | 3 (9.4) |
| T2, n (%) | 89 (17.9) | 0 (0) | 0 (0) | 86 (38.1) | 3 (13.6) | 0 (0) |
| T3, n (%) | 143 (28.8) | 0 (0) | 0 (0) | 139 (61.5) | 1 (4.5) | 3 (9.4) |
| T4, n (%) | 23 (4.6) | 0 (0) | 0 (0) | 0 (0) | 18 (81.8) | 5 (15.6) |
| T5, n (%) | 21 (4.2) | 0 (0) | 0 (0) | 0 (0) | 0 (0) | 21 (65.6) |

VTE, venous thromboembolism; FVL, factor V Leiden mutation; F2G20210A, prothrombin G20210A mutation; AT, antithrombin; PC, protein C; PS, protein S; APS, antiphospholipid syndrome; LA, lupus anticoagulant; aPL, antiphospholipid antibody

**Table 3.**
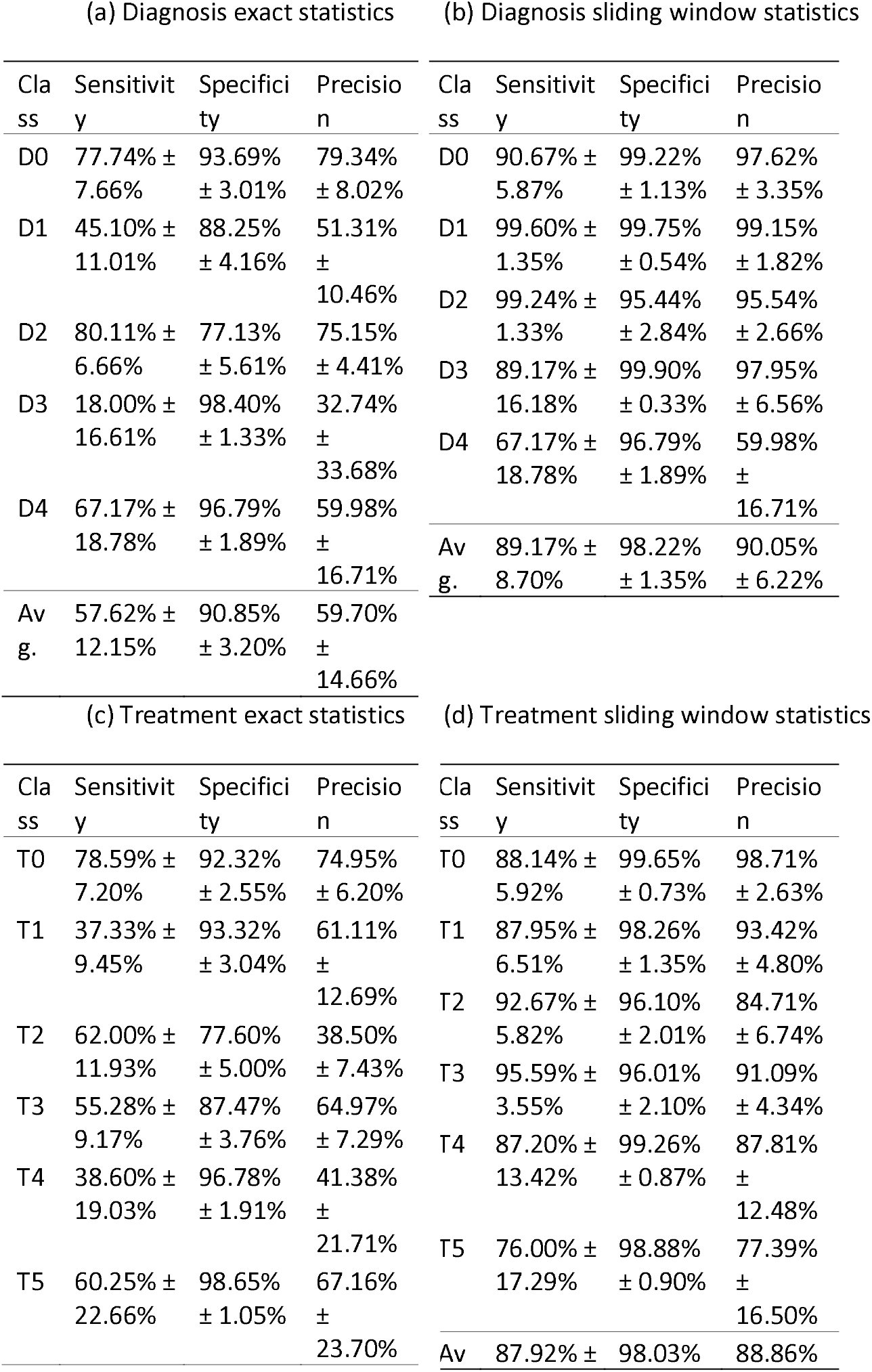
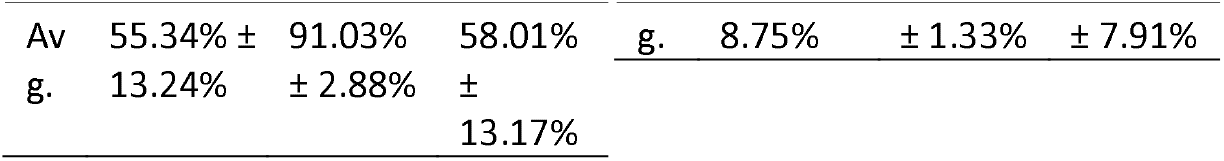
Mean ± std sensitivities, specificities, and precisions for diagnosis and treatment predictions. Tables 3b and 3d show the statistics with predictions treated as correct if within one ordinal class distance to the actual ordinal class. Since diagnosis D4 is categorical, the results do not change compared to those in Table 3a. Avg.: average.

### Classification Performance

In the diagnosis prediction, sensitivity under the exact evaluation was highest for class D2 (80.11% ± 6.66%), followed by D0 (77.74% ± 7.66%). Sensitivity was lower for D1 (45.10% ± 11.01%) and especially for D3 (18.00% ± 16.61%). The average sensitivity across all diagnosis classes was 57.62% ± 12.15%. With the sliding window approach, sensitivity improved above 89% across all ordinal classes. D1 reached the highest sensitivity with 99.60% ± 1.35%, and the other ordinal classes also showed high values, with D2 at 99.24% ± 1.33%, D0 at 90.67% ± 5.87%, and D3 at 89.17% ± 16.18%. The average sensitivity increased to 89.17% ± 8.70%.

Specificity for diagnosis prediction in the exact evaluation was generally strong, with values exceeding 77% for all classes. The highest specificity was observed for D3 (98.40% ± 1.33%) and D4 (96.79% ± 1.89%). The average specificity was 90.85% ± 3.20%. In the sliding window approach, specificity further improved, reaching above 95% for all classes. D3 again had the highest specificity (99.90% ± 0.33%), leading to an average specificity of 98.22% ± 1.35%.

Precision for diagnosis prediction in the exact evaluation was highest for D2 (75.15% ± 4.41%) and D0 (79.34% ± 8.02%). However, precision for D3 (32.74% ± 33.68%) showed high variability and was substantially lower. The average precision was 59.70% ± 14.66%. With the sliding window approach, precision rose above 95% for all ordinal classes. The average precision improved to 90.05% ± 6.22%.

In the treatment prediction, exact evaluation sensitivity varied greatly across classes. T0 achieved the highest sensitivity (78.59% ± 7.20%), while T1 and T4 lagged behind at 37.33% ± 9.45% and 38.60% ± 19.03%, respectively. The average sensitivity was 55.34% ± 13.24%. In the sliding window approach, T3 reached the highest sensitivity at 95.59% ± 3.55%, and the lowest was T5 with 76.00% ± 17.29%. The average sensitivity increased to 87.92% ± 8.75%.

Specificity in treatment prediction under exact evaluation was highest for T5 (98.65% ± 1.05%) and T4 (96.78% ± 1.91%). All classes exceeded 77%, resulting in an average specificity of 91.03% ± 2.88%. With the sliding window approach, specificity was above 96% for all classes. T0 had the highest specificity (99.65% ± 0.73%), and the average reached 98.03% ± 1.33%.

Precision in the exact treatment prediction showed more variability. T0 (74.95% ± 6.20%) received the highest value, while T2 received the lowest value (38.50% ± 7.43%). The average precision was 58.01% ± 13.17%. In the sliding window approach T0 achieved 98.71% ± 2.63%, and all other classes exceeded 77%, leading to a much higher average precision of 88.86% ± 7.91%.

### Correct Classification vs. Misclassification Patterns

For diagnosis prediction, the two-stage models correctly classified the majority of D0 (17.88 ± 1.76 out of 23), D1 (9.47 ± 2.31 out of 21), D2 (36.85 ± 3.06 out of 46), and D4 (4.03 ± 1.13 out of 6) samples. Misclassification rates increased for D1 and D3 (0.72 ± 0.66 out of 4 correctly classified), which showed overlapping responses with D2 (6.27 ± 2.10 out of 21 and 2.35 ± 1.03 out of 4, respectively) (see Figure 1a).

**Figure 1.**
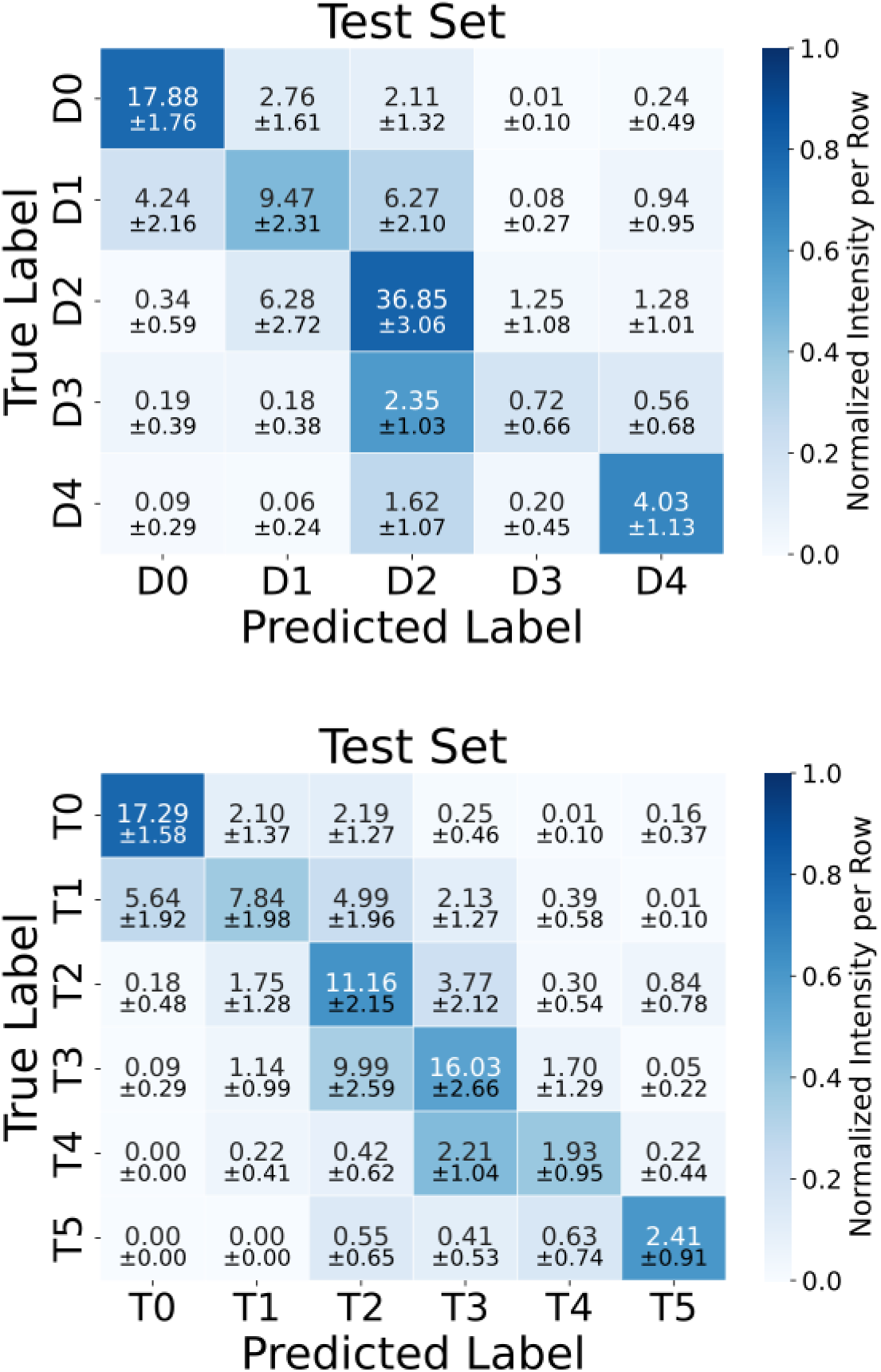
Confusion matrices of the two-stage model (diagnosis, D0-D4) and the ordinal model (treatment, T0-T5).

For treatment prediction, the ordinal model correctly classified the majority of T0 (17.29 ± 1.58 out of 22), T1 (7.84 ± 1.98 out of 21), T2 (11.16 ± 2.15 out of 18), T3 (16.03 ± 2.66 out of 29), and T5 (2.41 ± 0.91 out of 4), while for T4 only 1.93 ± 0.95 out of 5 were classified correctly (see Figure 1b).

### SHAP Value Analysis

For diagnosis, the most influential features in the first stage, based on absolute mean SHAP values, were the aPL index, anticardiolipin (aCL) antibody IgG, and aCL antibody IgM with values of 0.143, 0.051, and 0.030, respectively (Figure 2a). In the second stage, the most influential features were age at the first thrombosis, risk classification spontaneous thrombosis, and activated protein C resistance (APCR) with values of 0.507, 0.178, and 0.112, respectively (Figure 2b). Age at the first thrombosis was especially important for D0 and D2 (0.220 ± 0.016 and 0.201 ± 0.018), risk classification spontaneous thrombosis was especially important for D1 and D2 (0.063 ± 0.013 and 0.065 ± 0.013), and APCR was especially important for D0 and D1 (0.052 ± 0.008 and 0.045 ± 0.009).

**Figure 2.**
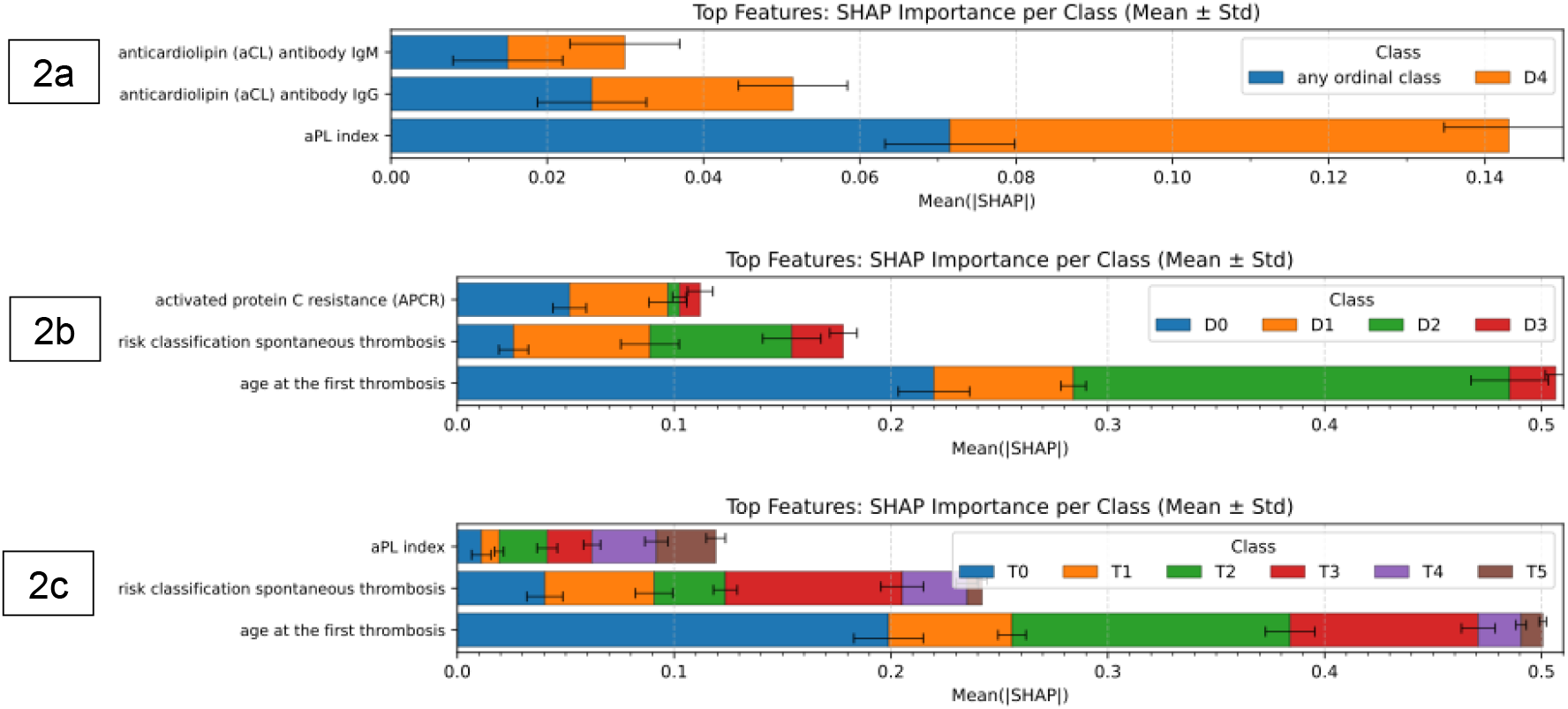
Top three features with the highest absolute mean ± standard deviation SHAP values per stage and target feature (diagnosis: 2a-b, treatment: 2c) of 100 runs with 100 different train and test splits

For treatment, the most influential features were age at the first thrombosis, risk classification spontaneous thrombosis, and aPL index (0.501, 0.242, and 0.119, respectively; Figure 2c). Age at the first thrombosis was especially important for T0, T2, and T3 (0.199 ± 0.016, 0.128 ± 0.011, and 0.087 ± 0.008), whereas risk classification spontaneous thrombosis was especially important for T3 (0.082 ± 0.010). The aPL index had the highest impact on T4 and T5 (0.030 ± 0.005 and 0.027 ± 0.004).

### Divergence of Model Predictions from Practitioner Predictions

The experiment was repeated 100 times with 100 unique and random train-test splits, and as a result each patient appeared approximately 20 times in the test set. For these runs, we assessed how well the algorithm could classify the patients on average. Of the 496 patients, 362 (73%) had no difference between the AI-based predictions and the practitioner diagnosis and subsequent treatment recommendations. For the remaining 134 patients, there was a divergence from the practitioner diagnosis in at least 20% of analyses for which the patient was part of the test set. The discrepancies are illustrated in the alluvial plot in Figure 3.

**Figure 3:**
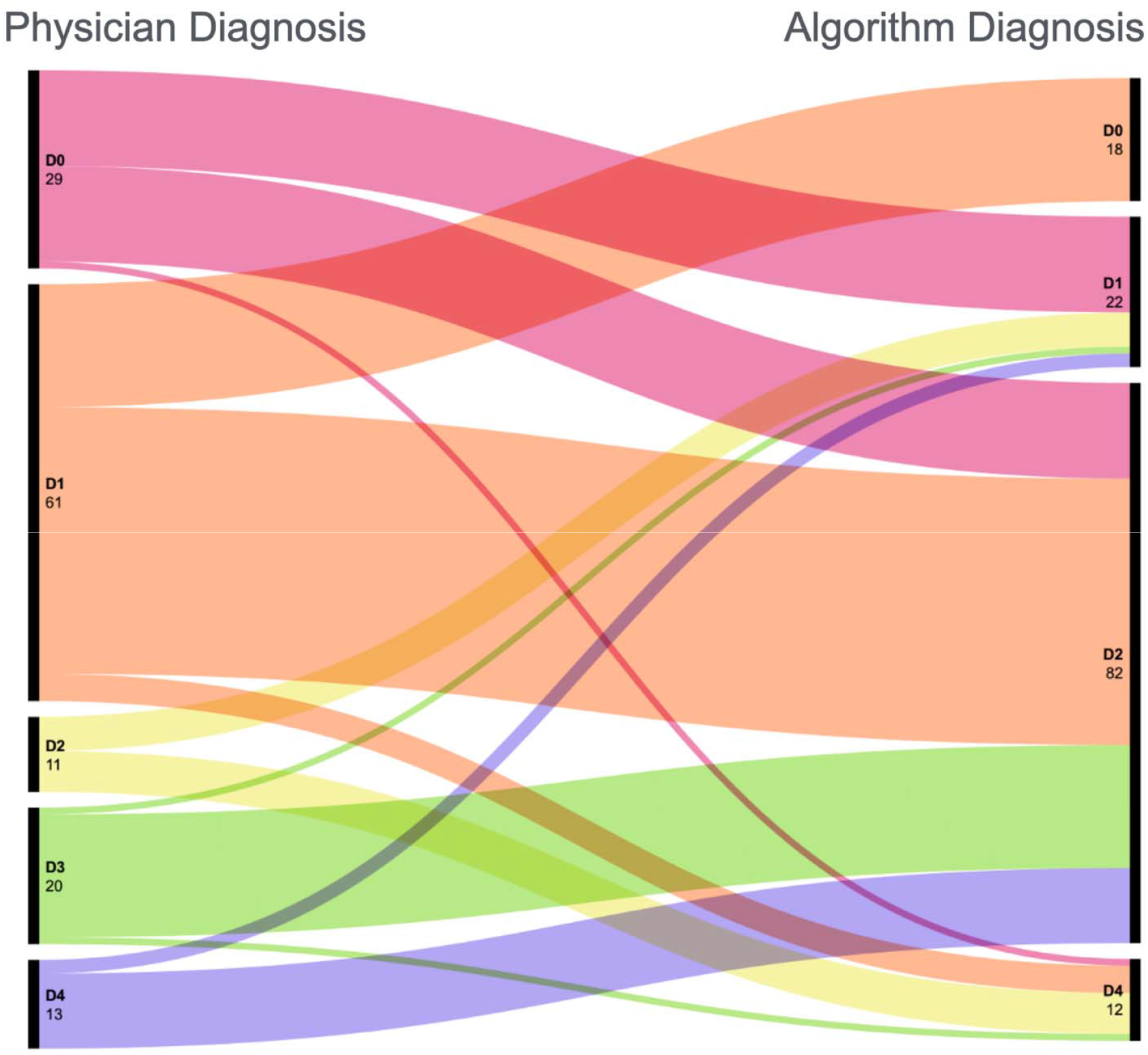
Discrepancies in diagnosis as given by the practitioner vs the algorithm. Alluvial plot (Sankey diagram) showing the absolute number of anomalous cases per diagnosis category (total n=134). The category D3 is omitted from the right-side axis because no cases were falsely given D3 by the algorithm.

Of the 29 cases in which thrombophilia was ruled out by the clinician, the algorithm classified 14 as low-risk thrombophilia, 14 as intermediate-risk thrombophilia, and one as APS. Re-evaluation of the patient data revealed that the APS patient, one intermediate-risk patient, and two low-risk patients had been misclassified by the clinician due to overlooked laboratory data. The remainder had been incorrectly classified by the algorithm.

Of the 61 cases classified as low-risk thrombophilia by the clinician, the algorithm classified 18 as thrombophilia excluded, 39 as intermediate-risk thrombophilia, and four as APS. Re-evaluation confirmed the thrombophilia excluded diagnosis in four of the 18 cases that the AI classified as thrombophilia excluded, while in the remaining cases, the AI slightly overestimated the thrombotic risk. The AI diagnosis was confirmed in 15 of the 39 cases that the AI-classified as intermediate-risk thrombophilia. One of the four AI-classified APS cases was misclassified by the physician due to an incomplete analysis of the aPL results. Interestingly, two of the 61 patients were found to have been incorrectly classified by both the clinician and the algorithm due to the presence of a high-risk thrombophilia diagnosis with a paradoxical clinical and laboratory presentation. One patient had AT-deficiency confirmed via genetic testing, but had borderline AT levels; however, the patient had experienced spontaneous DVT and PE at the age of 41, which confirmed a high-risk thrombophilia diagnosis. The second patient had double heterozygous FVL and F2G20210A, and experienced one instance of VTE in the setting of COC use at age of 29. Both of these cases were classified as high-risk thrombophilia upon re-evaluation.

Of the 11 cases diagnosed as intermediate-risk thrombophilia by the clinician, the algorithm correctly classified ten. The physician errors were attributed to an incomplete assessment of aPL results (n=6) or an incorrect interpretation of the patient history (n=4). One case was misclassified as low-risk thrombophilia by the algorithm.

Of the 20 cases diagnosed as high-risk thrombophilia by the physician, the algorithm classified one as low-risk, 18 as intermediate-risk, and one as APS. The algorithm was found to be incorrect in all cases except for the APS patient, where the physician had overlooked some of the aPL results, and one intermediate-risk patient, where it appeared to be a data-entry error by the physician. The algorithm underestimated the VTE risk in the remainder of cases.

Of the 13 cases that received an APS diagnosis from the physician, the algorithm classified 12 as intermediate-risk thrombophilia and one as low-risk thrombophilia. Re-evaluation of the patient records revealed that the patient AI-classified as low-risk thrombophilia was misclassified by the physician due to a data entry error. In the remaining 12 cases, the aPL results and the thrombophilia probability score were not correctly interpreted by the algorithm.

Overall, the AI correctly classified the patient in 36 out of 134 cases where there was a discrepancy between the AI and physician diagnoses. Of the remaining 98 patients, the algorithm underestimated the thrombophilia state in 71 cases and overestimated it in 27.

## Discussion

The diagnosis and management of thrombophilia is inherently complex, requiring nuanced interpretation of clinical and laboratory findings. This study demonstrated that a two-stage machine learning (ML) model can offer meaningful support in this context by accurately predicting thrombophilia diagnoses and treatment strategies using structured patient data. While the algorithm performed robustly across several domains, it also revealed key areas where human expertise and machine interpretation diverged. This highlights not only the potential but also the current limitations of AI-assisted decision-making in clinical hematology.

### ML Performance and Feature Importance

The integration of a two-stage model architecture—categorical classification followed by ordinal prediction—proved critical in navigating the hybrid nature of the diagnostic and treatment classes. The diagnosis of APS did not fall neatly within a linear progression and were thus modeled as a categorical variable. In contrast, ordinal categories like thrombophilia state (D0–D3) or strength of treatment recommendations (T0–T5) are inherently rankable, representing increasing disease severity or treatment intensity.

Feature importance and SHAP analyses underscored the clinical validity of model behavior. The APA results consistently emerged as the most influential feature in distinguishing categorical versus ordinal classes with the XGBClassifier models. This aligns with clinical understanding, as triple-positive aPL profiles (reflected in high APA index values) are strongly associated with APS and thus justify separate categorization. In distinguishing the ordinal classes with the XGBOrdinalV2 models, age at first thrombosis and spontaneous thrombosis were the most influential features, which corresponds with clinical practice, in which thrombosis at an earlier age and/or unprovoked thrombosis often indicate an underlying inherited risk. Other risk classification inputs and the number of thrombotic events also scored highly in the feature importance rankings. Their significance corroborates existing literature and reflects validated risk stratification frameworks that incorporate recurrence risk, inherited factors, and clinical history to guide both diagnostic categorization and treatment decisions [3, 5, 20, 25].

### Classification Accuracy and Use of the Sliding Window

Sensitivity, specificity, and precision analyses demonstrated high classification accuracy for the most prevalent categories—particularly D0 and D2 for diagnosis and T0 and T3 for treatment. These categories represent mild-to intermediate-risk thrombophilia with clear clinical thresholds and robust supporting data, which likely contributed to the model’s reliable performance.

However, the models showed notable underperformance in rarer categories, particularly D3 and T4, which were frequently misclassified. This is likely due to class imbalance, where these categories are underrepresented in the training data, which reduces the model’s ability to generalize predictions effectively. The implementation of a sliding window approach significantly improved performance metrics across ordinal classes, particularly for D3 and T4. For instance, D3 classification sensitivity rose from 18% to 89%. These gains highlight the model’s capacity to differentiate near-boundary cases when a tolerance buffer is applied, a method that could have clinical utility in prioritizing cases for secondary review.

### Algorithm vs. Clinician Judgment: Sources of Discrepancy

A key strength of this study lies in its comparative analysis between algorithmic predictions and clinician-assigned diagnoses. In 134 of 496 cases, the algorithm diverged from the clinician’s diagnosis in at least 20% of runs, warranting closer scrutiny.

In several cases, discrepancies were attributable to data entry errors or incomplete laboratory evaluation—human limitations which the algorithm helped uncover. For example, in 12 of the 13 cases where the clinician diagnosed D4 and the algorithm predicted D2, the algorithm was correct, indicating its ability to consistently interpret APA results according to standardized thresholds.

In contrast, the algorithm frequently underestimated risk in cases where patients had D3 diagnoses (e.g., confirmed AT deficiency or compound heterozygous FVL/F2G20210A) but with atypical laboratory profiles. This suggests that while the model learned associations between certain clinical markers and risk levels, it may have difficulty recognizing cases where functional impairment is present despite normal laboratory values—scenarios clinicians are trained to identify.

Misclassification of APS also occurred, especially when laboratory findings were robust (e.g., triple-positive APA) but associated clinical events were limited or ambiguous. This suggests that the model may underweight laboratory findings in the absence of corresponding clinical events. A model enhancement incorporating APS-specific criteria—such as revised Sapporo classification parameters— could help improve recognition of these cases [26].

Conversely, human decision-making appeared more prone to subjective risk interpretation. Several patients were assigned higher or lower risk classes by clinicians based on factors not included in the model, such as psychosocial elements, patient preference, or undocumented contextual clues (e.g., clinician intuition). While this underscores the value of human judgment, it also points to the potential benefit of an objective model to serve as a second-opinion tool, particularly in borderline or ambiguous cases.

### Interpretability and Clinical Utility

A notable strength of the two-stage model is its interpretability. By breaking down classification decisions into categorical versus ordinal tasks, the model offers a structure that mirrors clinical logic. Furthermore, the inclusion of feature importance metrics provides transparency—an essential factor in clinical implementation. For example, understanding that APA index or age at first thrombosis drove a specific prediction enables clinicians to weigh model outputs alongside their own assessments and interpret algorithmic findings within the clinical context.

In clinical practice, such a model could function as a decision support tool: flagging high-risk cases, recommending further testing, or validating the physician’s interpretation. It could also help standardize treatment decisions in settings where specialist expertise is lacking, offering tailored anticoagulation guidance based on reproducible criteria.

### Limitations and Future Directions

Despite these findings, several limitations should be considered. First, the model was trained and validated using data from a single center, which raises concerns about generalizability. External validation in prospective studies across diverse populations and care settings is essential prior to implementation in routine clinical practice. Second, class imbalance affected model accuracy, particularly in rare classes. Addressing this issue through data augmentation, synthetic resampling techniques, or alternative model architectures may further enhance predictive performance. Third, while the model included a wide range of laboratory and clinical variables, several relevant contextual features were not assessed—notably imaging results, comorbidities, and patient-specific treatment preferences. Future iterations could integrate natural language processing (NLP) of EMRs or unstructured physician notes to capture these contextual nuances. Lastly, the study’s retrospective design inherently limits causal interpretation. While the models predict diagnoses and treatment based on existing data, they cannot capture evolving clinical reasoning or decision-making in real time. Prospective trials are needed to evaluate whether AI-supported decisions improve patient outcomes and workflow efficiency.

## Conclusion

This study presents a novel two-stage ML approach that successfully classifies thrombophilia diagnosis and treatment strategy with promising accuracy and interpretability. By employing a two-stage approach with XGBClassifier and XGBOrdinalV2 to diagnosis and XGBOrdinal to treatment, we successfully predicted categorical and ordinal classifications with high specificity and moderate sensitivity and precision. The implementation of a sliding window approach further improved classification performance for ordinal categories. Future research should focus on refining these models through external validation, prospective studies, and integration with clinical decision-support systems to improve thrombophilia management and patient outcomes. These findings contribute to the growing body of evidence supporting the use of ML in complex medical decision-making processes.

## Data Availability

All data produced in the present study are available upon reasonable request to the authors.

## Acknowledgements

We thank the clinical coagulation laboratory of the University Hospital Bonn for their technical expertise.

## Author Contributions

BP conceived and supervised the study. HLM, BP, and HR collected and analyzed the data. FK, MK, and SMJ developed the machine learning pipeline. HLM and FK wrote the manuscript. SJM and BP edited the manuscript. All authors reviewed and approved the final version of the manuscript.

## Conflict of Interest

The authors do not have any relevant conflicts of interest to declare.

## Supplemental Material

**Supplemental Figure S1:**
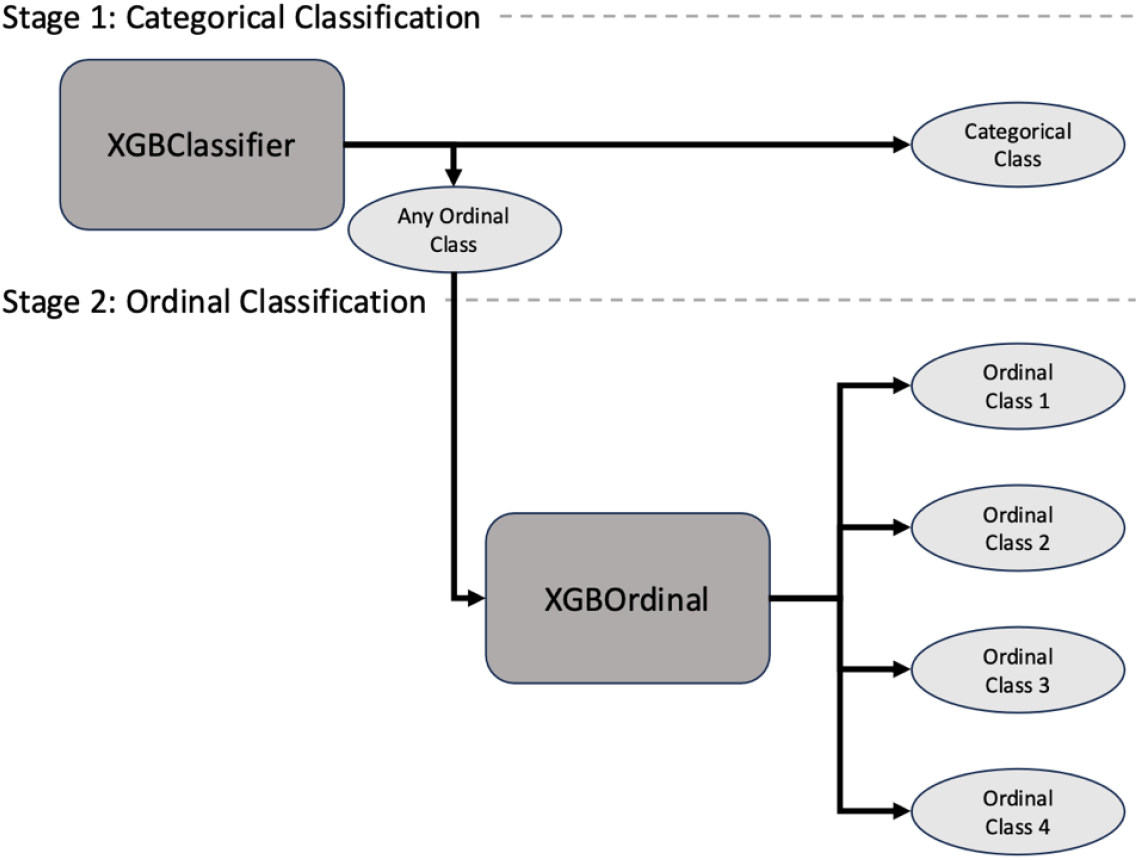
Two-stage approach to classification of the dataset

### Preprocessing

Data preprocessing was conducted to make the data compatible with the algorithms, including replacing typos, dropping unnecessary features, and applying one-hot encoding to the ten categorical features. One-hot encoding transforms categorical data into separate columns for each unique category assigning a 1 to the column that matches the class for a given row and a 0 to all others (Supplemental Figure S2, patients A and B). For entries containing multiple classes separated by commas, separate binary columns were created for each class, and all were set to 1. For instance, a value of “risk evaluation, familial thrombophilia” in the feature “indication” was transformed into that patient having “risk evaluation” and “familial thrombophilia” (Supplemental Figure S2, patient C). This led to 119 numerical features.

**Supplemental Figure S2:**
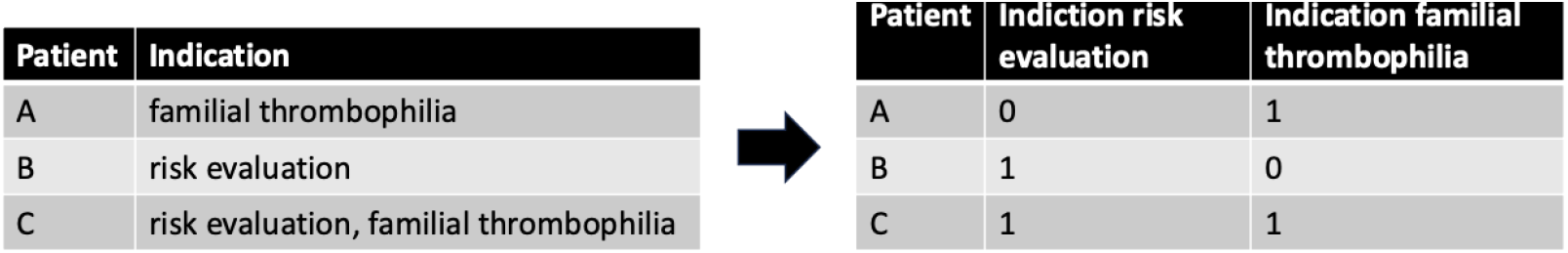
Patients A and B show the standard one-hot encoder and patient C shows the comma-separated one-hot encoder.

### Technical Details

For each target feature, five unique two-stage models with random stratified train-test-splits with a test set size of 20% (stratified on respective target feature) were trained. Per two-stage model, the same train-test-split for training and testing of the underlying XGBClassifier and XGBOrdinalV2 models was used. The default hyperparameters were used and hyperparameter tuning was not performed. Imputing missing values was unnecessary since XGBClassifier and XGBOrdinalV2 can handle missing values. Feature scaling was also not required since XGBClassifier and XGBOrdinalV2 are based on decision trees, which are not sensitive to the scale of input features.

## Notes

### Competing Interest Statement

The authors have declared no competing interest.

### Funding Statement

This study did not receive any funding.

### Author Declarations

The Ethics Committee of the University of Bonn Faculty of Medicine sanctioned the collection of data from the electronic medical record and waived the requirement for written informed consent due to the retrospective nature of the study.

